# Integrating Plasma pTau-217 and Digital Cognitive Assessments for Early Detection in Alzheimer’s Disease

**DOI:** 10.1101/2025.03.03.25323297

**Authors:** Casey R. Vanderlip, Craig E.L. Stark

**Affiliations:** Department of Neurobiology and Behavior, 1424 Biological Sciences III Irvine, University of California Irvine, Irvine, CA, 92697 USA

**Keywords:** pTau217, Digital cognitive assessments, Alzheimer’s disease, memory, low-burden measures, early detection

## Abstract

Plasma pTau-217 has emerged as a sensitive and specific biomarker for early Alzheimer’s disease detection. However, the timeline of pathological changes and the onset of cognitive decline remain unclear. On the other hand, digital cognitive assessments have also shown promise in detecting subtle cognitive changes, but the sensitivity and specificity of these assessments is not fully understood. Here, we investigate whether combining these low-burden tools can improve the identification of cognitively unimpaired individuals at high risk for future cognitive decline. We analyzed 954 amyloid-positive cognitively unimpaired individuals who completed a brief digital cognitive assessment and a blood test for pTau-217, evaluating their ability to identify those at high risk for decline on the Preclinical Alzheimer’s Cognitive Composite (PACC) and the Mini-Mental State Exam (MMSE). Further, we investigated whether the predictive value of these measures differed by sex or APOE status. We found that combining memory performance with pTau-217 enhanced the ability to identify individuals who declined on the PACC and MMSE over the next five years, even after controlling for age, sex, education, and baseline cognitive performance. Specifically, individuals with both elevated pTau-217 and low memory performance were at a greater risk for future decline than those with either risk factor alone. Notably, the predictive value of these measures did not differ by sex but was significantly stronger in APOE4 noncarriers compared to carriers. Together, this suggests that combining a brief digital cognitive assessment with plasma pTau-217 provides a reliable and sensitive method for identifying individuals at high risk for future cognitive decline in Alzheimer’s disease.

## 1. Introduction

Alzheimer’s disease (AD) is a progressive neurodegenerative disorder and the leading cause of dementia worldwide. It is characterized by the accumulation of amyloid-beta (Aβ) plaques and neurofibrillary tangles composed of hyperphosphorylated tau, developing gradually over decades before clinical symptoms emerge^1–3^. The earliest phase, known as the preclinical stage, involves subtle biological and cognitive changes that often go unnoticed in standard clinical assessments^4–6^. While early detection of individuals with elevated AD pathologies is crucial for timely intervention, not everyone with AD pathology follows the same cognitive trajectory. Some individuals remain cognitively stable for decades despite pathological changes, making it difficult to determine when intervention is necessary^7^. This variability in disease progression poses a significant challenge in predicting who will develop AD-related impairment and when cognitive decline will occur.

Recent advances in fluid biomarkers have significantly improved the ability to detect AD pathology before cognitive symptoms appear^8–10^. Among these, plasma pTau-217 has emerged as a highly specific and sensitive marker of amyloid pathology and disease progression. Studies have shown that elevated plasma pTau-217 levels strongly correlate with amyloid burden in the brain, making it a powerful tool for early detection^11,12^. Notably, recent guidelines suggest that pTau-217 is the only plasma biomarker recommended for AD diagnosis^1^. However, while pTau-217 reliably indicates underlying pathology, it does not directly measure cognitive function, creating uncertainty about its ability to predict when an individual will experience cognitive decline.

Cognitive assessments, particularly digital cognitive tests, offer a complementary approach to biomarker-driven detection by capturing subtle functional changes that may signal early AD-related cognitive decline. Digital assessments are scalable, cost-effective, and sensitive to early cognitive changes that might be missed by traditional paper-based tests^13–17^. In particular, digital cognitive tests that tax hippocampal memory have proven to be highly sensitive to early AD-related changes, including detecting amyloid and tau pathology and predicting future clinical symptoms^18–20^. Some evidence even suggests that these assessments may outperform biological biomarkers in identifying individuals at high risk for future cognitive decline^21^. However, the specificity of these cognitive measures in distinguishing AD-related cognitive changes from normal aging or other conditions remains unclear.

Given the distinct strengths of fluid biomarkers and cognitive assessments, an integrated approach combining plasma pTau-217 with a digital cognitive battery may enhance the ability to identify cognitively unimpaired individuals at high risk for future decline. By leveraging both biological markers of AD pathology and sensitive cognitive performance metrics, this approach has the potential to improve early detection and risk stratification.

In this study, we examined whether combining plasma pTau-217 with a brief digital cognitive battery consisting of three hippocampal-based memory tasks could reliably identify cognitively unimpaired individuals at high risk for future cognitive decline. Specifically, we hypothesized that combining pTau-217 levels with digital cognitive performance would improve the accuracy and reliability of identifying individuals at risk for cognitive decline over the next five years. We analyzed data from nearly 1,000 cognitively unimpaired individuals in the A4 clinical trial who underwent both a blood test for pTau-217 and a digital cognitive assessment at baseline. We assessed the predictive ability of these measures individually and in combination to predict future decline on established cognitive assessments, including the Preclinical Alzheimer’s Cognitive Composite (PACC) and the Mini-Mental State Exam (MMSE). Additionally, we examined whether their predictive value differed by sex or APOE genotype, well-established AD-related risk factors.

By assessing both biological and cognitive markers, this study aims to refine early detection strategies for AD, providing a more reliable and sensitive method for identifying at-risk individuals. If successful, this approach could enhance clinical decision-making, inform early interventions, and improve trial recruitment for disease-modifying therapies.

## 2. Methods

All data were obtained from the A4 clinical trial, a study whose design and aims have been described in detail previously^22–24^. Briefly, A4 was a double-blind, placebo-controlled, 240-week Phase 3 trial evaluating an anti-Aβ monoclonal antibody in cognitively unimpaired older adults with preclinical AD. All participants (n = 954) were Aβ-positive based on florbetapir PET scan results, aged 65 to 85, and classified as cognitively unimpaired, with an MMSE score above 24 and a Global Clinical Dementia Rating (CDR) score of 0 (Table 1).

**Table 1:** Demographics.

|  | Carrier | Noncarrier | 89 |
| --- | --- | --- | --- |
| n | 594 | 360 |  |
| Age | 71.30 (4.51) | 73.26 (5.03) |  |
| Sex (% Female) | 42.10% | 38.30% |  |
| Education | 16.64 (2.71) | 16.54 (2.95) |  |

### 2.1. Participants

All data were obtained from the A4 clinical trial, a study whose design and aims have been previously described in detail. Briefly, A4 was a double-blind, placebo-controlled, 240-week Phase 3 trial evaluating an anti-Aβ monoclonal antibody in cognitively unimpaired older adults with preclinical AD. The study included 954 Aβ-positive participants, as determined by florbetapir PET scan results, aged 65 to 85, with an MMSE score above 24 and a Global Clinical Dementia Rating (CDR) score of 0. Additionally, each participant completed a brief digital cognitive battery at screening and underwent a blood test at baseline.

### 2.2. Digital Cognitive Battery

The Computerized Cognitive Composite (C3) is a brief digital cognitive battery completed by all A4 participants. This composite has been previously described in detail^19,20^. Briefly, it includes three hippocampal memory tasks, the Behavioral Pattern Separation Task (BPST), Face-Name Task (FNAME), and One Card Learning Task (OCL), along with three control tasks: a detection psychomotor speed task, an identification visual attention task, and a one-back working memory task. Prior research has shown that hippocampal memory-based tasks are more sensitive to AD-related changes. Therefore, we focused our analyses on the three hippocampal memory tasks.

The BPSO, now known as the Mnemonic Similarity Task (MST), is a hippocampal memory task designed to assess pattern separation^25,26^. In this study, a shortened version of the MST was used. Briefly, during the encoding phase, participants viewed 40 images of everyday objects on a white background, making indoor/outdoor judgments via button press (5 s per image, 0.5 s inter-stimulus interval (ISI)). Immediately afterward, they received instructions for a recognition memory test, in which they classified objects as “old” (identical to a previously seen image), “similar” (a slightly altered version of a studied item, such as a different exemplar or rotation), or “new.” During this phase, participants viewed 60 images (5 s per image, 0.5 s ISI), consisting of 20 exact repeats from encoding (targets), 20 completely novel images (foils), and 20 similar but non-identical images (lures). The primary measure of interest was the lure discrimination index (LDI), calculated as the proportion of “Similar” responses to lures minus the proportion of “Similar” responses to foils, adjusting for response bias.

For the FNAME, Participants viewed 12 face-name pairs presented sequentially and judged whether each name “fit” the face to maintain attentiveness. They had 5 seconds to respond and were instructed to remember the pairings. After a 12- to 15-minute delay filled with other cognitive tasks, memory was assessed through three tasks: face recognition (FSBT), first-letter name recall (FNLT), and face-name matching (FNMT). In FSBT, participants identified previously learned faces from a set of three, including two distractors matched for age, race, and sex. In FNLT, they selected the first letter of the paired name using an on-screen keyboard. In FNMT, they chose the correct name from three options: the target name, a re-paired name, and a matched foil. Each task was scored out of 12, with FNMT accuracy serving as the primary outcome measure.

OCL is modeled off the MST and is a task that taxes hippocampal pattern separation which is critical for episodic memory. In this task, participants are shown a series of playing cards and are asked if they have seen the playing card previously during the task. Four cards are randomly selected to repeat eight times throughout the task. The task consists of 80 trials and the performance outcome is accuracy.

The C3 consisted of one primary outcome from each of the three memory tasks and is calculated as the average of these z-scored outcomes, standardized based on screening data.

### 2.3. Neuropsychological Assessments

All participants completed in-person, gold-standard neuropsychological testing, including the Preclinical Alzheimer’s Cognitive Composite (PACC)^27,28^. The PACC comprises four components: the total score on the Free and Cued Selective Reminding Test (FCSRT), delayed paragraph recall from the Logical Memory IIa test (Wechsler Memory Scale), the Digit Symbol Substitution Test (Wechsler Adult Intelligence Scale–Revised), and the MMSE total score. To minimize practice effects, alternate versions of these tests were used at each session. To control for baseline performance, each component score was converted to a z-score by subtracting the baseline mean and dividing by the baseline standard deviation. The PACC score was calculated as the sum of these z-scores, with negative values indicating cognitive decline.

### 2.4. Plasma pTau-217

Plasma samples were provided to Lilly to aid in developing a pTau-217 assay for cognitively impaired individuals. The Lilly Clinical Diagnostics Laboratory measured pTau-217 levels using an electrochemiluminescent immunoassay, with sample preparation automated by the Tecan Fluent workstation and detection conducted on the MSD Sector S Imager 600MM.

### 2.5. Statistical analyses

All analyses were conducted in RStudio. For baseline comparisons, low performers were defined as individuals scoring more than 1.5 standard deviations below the group average. To predict future cognitive decline, participants were categorized as progressors or non-progressors based on changes in the PACC or MMSE. At each timepoint, progressors on the PACC were defined as those who declined more than 1.5 standard deviations below their baseline performance relative to the group average. For MMSE analyses, a score below 25 was used as the cutoff for cognitive impairment, a commonly accepted clinical threshold.

Logistic regression models were used to predict which individuals would exhibit cognitive decline on either the PACC or MMSE. For all models, we used pTau217 at baseline and C3 scores from screening and both measures were z-scored to standardize values. To prevent overfitting and improve generalizability, all models were evaluated using leave-one-out (LOO) cross-validation. Model performance was assessed using area under the curve (AUC) values derived from receiver operating characteristic (ROC) curves. Confidence intervals were generated via bootstrapping with 1,000 permutations. To assess model sensitivity, true positive rates were calculated at a predefined false positive rate of 20% using the 1,000 bootstrap samples.

For odds ratio analyses, we examined the likelihood of future cognitive decline in individuals with either (1) pTau-217 levels one standard deviation above the group mean, (2) a C3 composite score one standard deviation below the mean, or (3) both risk factors. Z-tests were used to compare the differences in log odds across these conditions. Linear mixed effects models were

used to investigate longitudinal changes on the PACC and MMSE. Additionally, one-way ANOVAs with Tukey’s HSD post-hoc tests were performed to identify within-factor differences. For all analyses, p-values below 0.05 were considered statistically reliable.

## 3. Results

### 3.1. Only C3 Scores Differentiate High and Low PACC Performers at Baseline

To assess whether combining plasma pTau-217 and C3 scores enhances clinical utility, we began by initially examining their ability to predict PACC performance at baseline before turning to their ability to predict cognitive decline from baseline after either approximately two or five years. To investigate baseline performance, individuals were categorized as high or low performers on the PACC, with low performers defined as those scoring more than 1.5 standard deviations below the group average. We then trained four separate logistic regression models to predict high versus low performers (Fig 1A, top).

**Figure 1:**
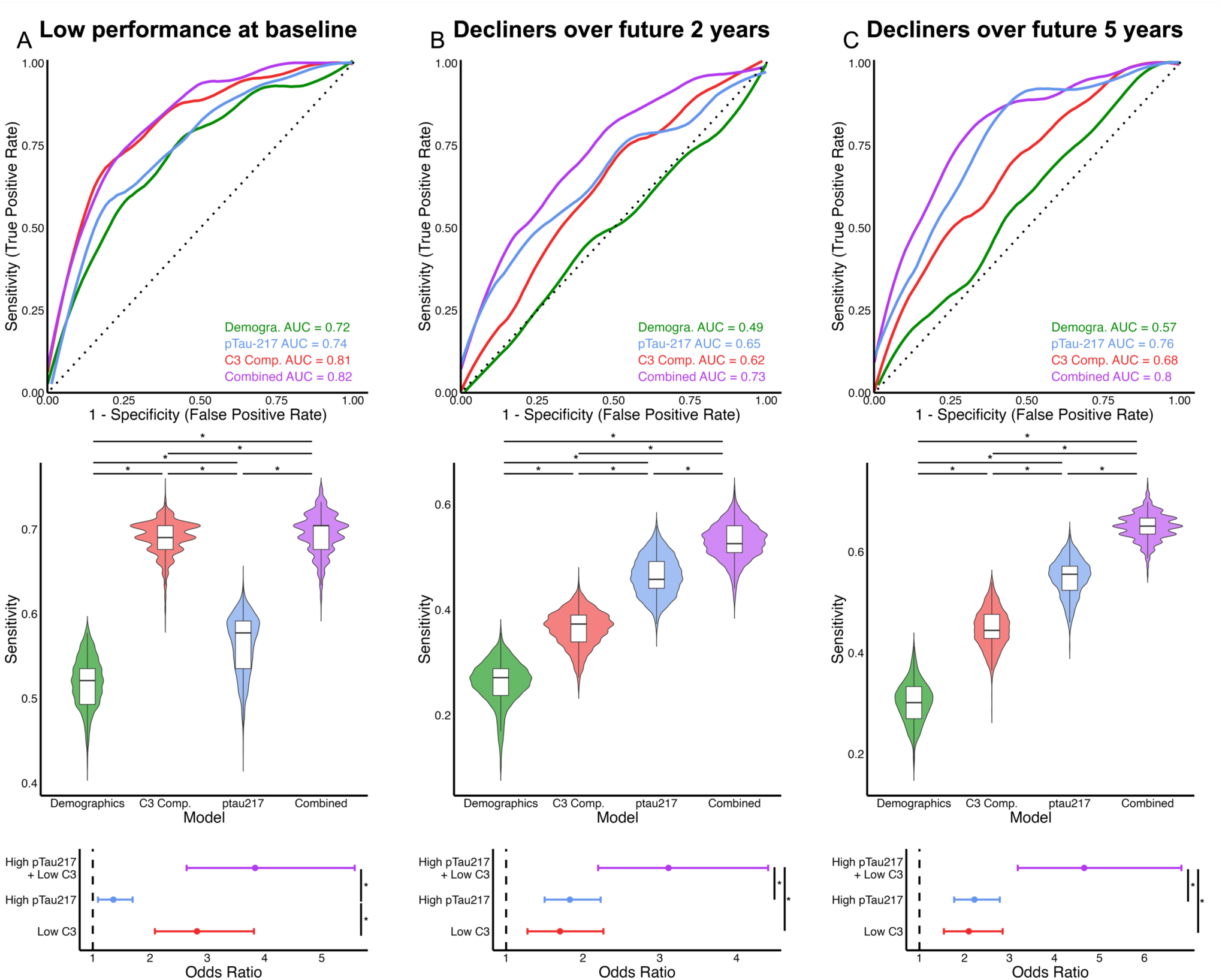
Integrating plasma pTau-217 and the C3 enhances prediction of future decline on the PACC. Comparison of models using demographics alone (green), demographics plus pTau-217 (blue), C3 composite (red), or both pTau-217 and C3 (purple) for predicting low performers and future cognitive decline. (Top) ROC curves illustrating the predictive value of each model. (Middle) Violin plots depicting model sensitivity at a false positive rate of 20%. (Bottom) Odds ratios for future decline based on low memory (red), high pTau-217 (blue), or both (purple). (A) At baseline, the C3 composite was the strongest predictor of high and low performers, showing the highest sensitivity and odds ratios. Over two (B) and five (C) years, the combined model (pTau-217 + C3) was the most predictive of future decline, achieving the highest AUCs and sensitivity. Critically, individuals with both high baseline pTau-217 and low baseline C3 performance had significantly higher odds of cognitive decline compared to those with only one of these risk factors.

The base model, which included demographic variables (age, sex, and education), reliably distinguished between high and low performers (LOO AUC: 0.716, CI: 0.651 – 0.778). Notably, adding pTau-217 to the demographic model did not improve predictive accuracy, suggesting that pTau-217 does not contribute additional explanatory power (LOO AUC: 0.741, CI: 0.679 – 0.796). In contrast, incorporating C3 performance into the demographic model significantly enhanced its predictive value for distinguishing low and high performers suggesting that C3 and PACC performance are correlated (LOO AUC: 0.811, CI: 0.755 – 0.861). Finally, a model combining demographics, C3 performance, and pTau-217 performed similarly to the model including only C3 performance, significantly outperforming the baseline demographic model but not adding substantial predictive benefit over C3 alone (LOO AUC: 0.822, CI: 0.776 – 0.863).

To further evaluate model sensitivity, we set an a priori false positive rate of 20% and calculated sensitivity across all bootstrapped models. Sensitivity values were compared across models, revealing significant differences. The C3 composite model outperformed all others, followed by the combined model, which performed better than the pTau-217 and base demographic models (Fig 1A, middle, Two-way ANOVAs, F(3, 3996) = 11,981, p < .0001, Tukey’s HSD: all ps < 0.001). Additionally, the pTau-217 model demonstrated greater sensitivity than the base model (p < 0.001). Lastly, we calculated the odds of low memory performance based on elevated pTau-217 (≥1 SD above the group mean), low C3 composite scores (≥1 SD below the mean), or both. Elevated pTau-217 was associated with a 36% increased likelihood of being in the low-performance group (Fig 1A, bottom OR = 1.360, CI = 2.085–3.815). In contrast, low C3 memory performance was linked to a significantly higher 182% increased risk (OR = 2.820, CI = 2.085–3.815), while having both elevated pTau-217 and low memory was associated with a 284% increased risk (OR = 3.835, CI = 2.639–5.575).

Z-tests confirmed that both low memory and the combined condition were significantly associated with greater odds of low performance, while the combined condition did not differ significantly from low memory alone (High pTau-217 vs. Low Memory: Z = −3.822, p < 0.001; Both vs. High pTau-217: Z = 4.679, p < 0.001; Both vs. Low Memory: Z = 1.253, p = 0.210). These findings suggest that, unsurprisingly, baseline cognitive performance is more strongly predicted by a digital cognitive test performance than to plasma pTau-217 levels.

### 3.2. Combining plasma pTau-217 and C3 scores enhance prediction of future cognitive decline

It is unsurprising that C3 performance was a stronger predictor of high and low PACC performers compared to pTau-217, as both the C3 and PACC are cognitive measures designed to assess early stages of dementia. However, the true test of its utility lies in its ability at baseline or screening to predict future decline on the PACC. Thus, we examined whether these measures could predict individuals who experienced the greatest cognitive decline over 96 weeks (∼1.9 years). Decline was calculated as the difference between PACC scores at the 96-week visit and baseline, thus baseline performance was controlled for. Demographics alone were not sufficient to identify those who exhibited the most decline (Fig 1B, top LOO AUC: 0.490, CI: 0.472 – 0.588). However, adding either pTau-217 or C3 performance individually improved predictive accuracy above chance, with both models reaching similar AUCs (pTau-217: LOO AUC: 0.654, CI: 0.571 – 0.733, C3: LOO AUC: 0.622, CI: 0.557 – 0.695). Notably, the best predictive model included both pTau-217 and C3 performance alongside demographics, suggesting that each measure provides complementary explanatory power (LOO AUC: 0.725, CI: 0.650 – 0.795). Furthermore, at the predefined false positive rate of 20%, the combined model demonstrated the highest sensitivity, outperforming all other models (Fig 1B, middle, Two-way ANOVA, F(3, 3996) = 10475, p < .0001, Tukey’s HSD: all ps < 0.001).

Examining odds ratios for cognitive decline over two years, we found that elevated baseline pTau-217 and low baseline memory performance were associated with an 83% and 70% increased risk of decline, respectively (Fig 1B, bottom pTau-217: OR = 1.830, CI = 1.501–2.230; Low memory: OR = 1.702, CI = 1.278–2.267). These odds did not significantly differ (Z = 0.408, p = 0.683). However, individuals with both high pTau-217 and low memory performance at baseline had a 211% increased risk of two-year decline (OR = 3.114, CI = 2.198–4.411). Importantly, the combined condition conferred a significantly greater risk of decline than either measure alone (Combined vs. pTau-217: Z = 2.601, p = 0.009; Combined vs. Low memory: Z = 2.626, p = 0.009). Together, these findings support the value of combining hippocampal memory performance and plasma pTau-217 to improve the prediction of future cognitive decline.

Next, we extended this analysis to examine whether baseline measures could predict cognitive decline over 240 weeks (∼4.6 years) and observed similar results. Once again, the most accurate model included both pTau-217 and C3 performance, reinforcing that these measures together improve the prediction of cognitive decline for up to five years (Fig 1C, top Demographics: LOO AUC: 0.570, CI: 0.485 – 0. 645, pTau-217: LOO AUC: 0.762, CI: 0.705 – 0.821, C3: LOO AUC: 0.679, CI: 0.612– 0.745, Combined: LOO AUC: 0.799, CI: 0.742–0.855). At the predefined false positive rate, the combined model demonstrated the highest sensitivity, followed by the plasma pTau-217 model (Fig 1C, middle, F(3, 3996) = 17171, p < .0001, Tukey’s HSD: all ps < 0.001). Examining odds ratios, we found that elevated pTau-217 was associated with a 122% increased risk of decline (Fig 1C, bottom OR = 2.222, CI = 1.772– 2.786), while low memory performance was associated with a 110% increased risk (OR = 2.097, CI = 1.544–2.850). These risks did not significantly differ (Z = 0.297, p = 0.767). However, individuals with both high pTau-217 and low memory performance at baseline had a 366% increased risk of decline (OR = 4.661, CI = 3.185–6.821), which was significantly greater than either measure alone (Combined vs. pTau-217: Z = 3.278, p = 0.001; Combined vs. Low memory: Z = 3.201, p = 0.001). These findings further support the utility of combining plasma pTau-217 and a brief cognitive battery to identify individuals at risk for future cognitive decline.

### 3.3. Clinical utility of combining plasma pTau-217 and a remote cognitive battery differs by APOE4 genotype but not by sex

AD risk differs by sex and APOE genotype, and the efficacy of currently approved treatments varies based on these risk factors^29–31^. Therefore, it is critical to determine whether the clinical utility of plasma pTau-217 and cognitive measures differs across subgroups. To investigate this, we conducted post hoc analyses, evaluating the ability of the combined model derived above (demographics, pTau-217, and C3 composite scores) to predict low versus high performers at baseline and cognitive decline over 96 and 240 weeks, stratified by sex and APOE genotype.

At baseline, model performance was similar for distinguishing low versus high performers across sexes, though it performed slightly better in females (Fig 2A, top, Males: LOO AUC: 0.773, CI: 0.700 – 0.841, Females: LOO AUC: 0.839, CI: 0.767 – 0.905). Likewise, we did not observe substantial differences in predictive accuracy by APOE4 carriership (Fig 2A, bottom, Noncarriers: LOO AUC: 0.856, CI: 0.793– 0.912, Carriers: LOO AUC: 0.801, CI: 0.738 – 0.860), suggesting that differentiating low and high performers does not differ by these risk factors.

**Figure 2:**
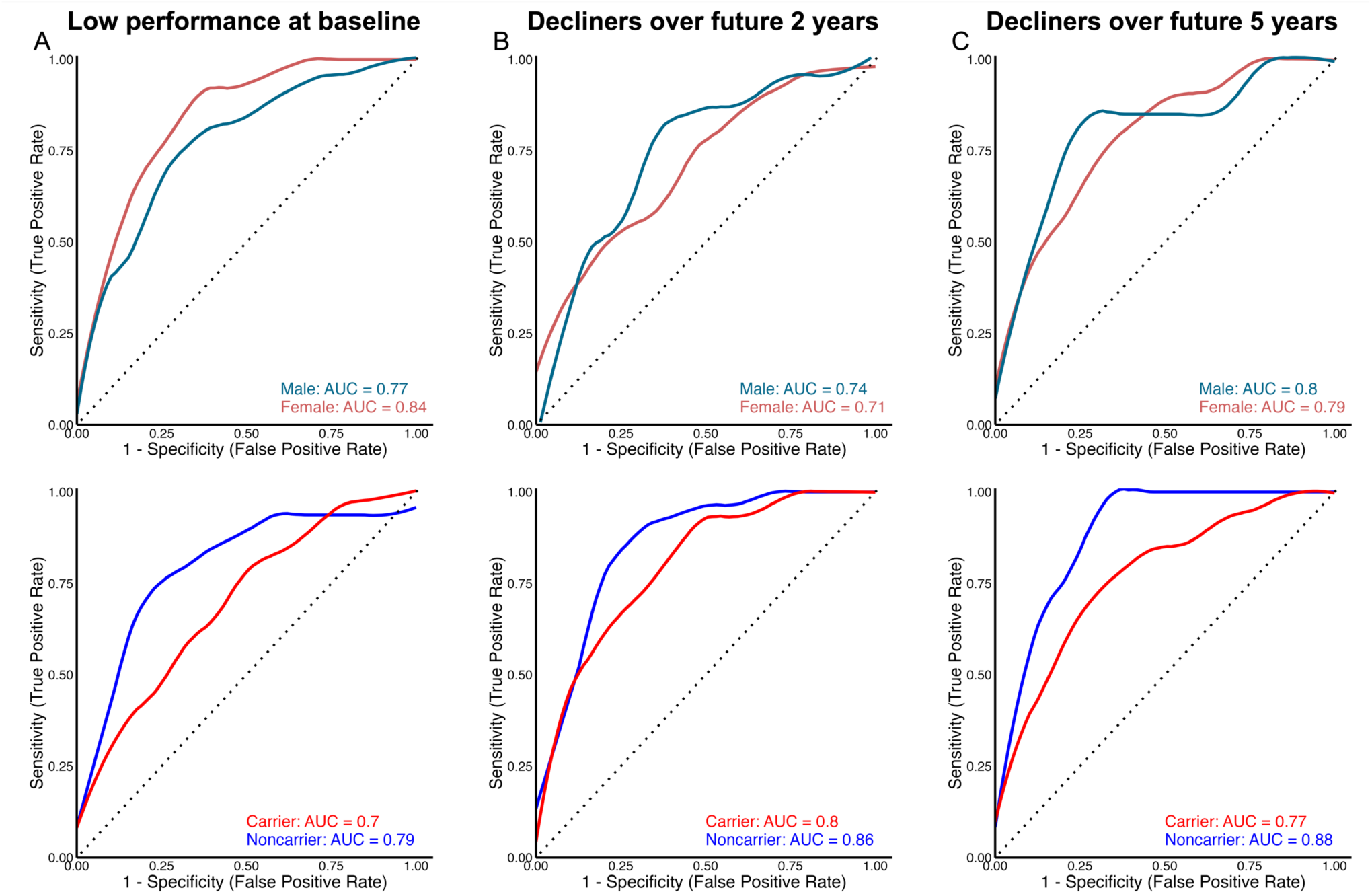
Predictive capacity of the combined model (demographics, plasma pTau-217 and C3) differs by APOE4 carrier status but not sex (Top) ROC curves illustrating the predictive value of the combined model in males (blue) and females (red). (Bottom) ROC curves illustrating the predictive value of the combined model in APOE4 noncarriers (blue) and APOE4 carriers (red). (A) At baseline, the model performed similarly in males and females, with a slight advantage in females (top). Additionally, no significant difference was observed between APOE4 carriers and noncarriers in baseline performance. (B–C) When predicting future cognitive decline, the models performed equally well in males and females at both two (B) and five (C) years. However, the predictive capacity was stronger in APOE4 noncarriers compared to carriers at both time points.

Examining prediction of cognitive decline over 96 and 240 weeks, we found no significant sex differences, indicating that the clinical utility of pTau-217 and the cognitive battery did not vary substantially by sex (Fig 2B, top, 2 years: Males: LOO AUC: 0.744, CI: 0.637 – 0.837, Females: LOO AUC: 0.712, CI: 0.615– 0.799, Fig 2C, top, 5 years: Males: LOO AUC: 0.804, CI: 0.681 – 0.903, Females: LOO AUC: 0.788, CI: 0.719 – 0.856). However, the models performed significantly better at predicting decline in APOE4 noncarriers compared to carriers (Fig 2B, bottom, 2 years: Noncarriers: LOO AUC: 0.794, CI: 0.654– 0.906, Carriers: LOO AUC: 0.696, CI: 0.621– 0.769, Fig 2C, bottom, 5 years: Noncarriers: LOO AUC: 0.885, CI: 0.821– 0.937, Carriers: LOO AUC: 0.772, CI: 0.692– 0.836), suggesting that APOE4 carriership may moderate the predictive value of these measures and should be APOE4 carriers may require different predictive markers or more tailored risk models to optimize early detection strategies.

### 3.4. Plasma pTau-217 and the C3 composite predict future impairment on the MMSE

The PACC is a well-established neuropsychological battery, but it is not commonly used in clinical settings, where global screening tools such as the MMSE are more prevalent. Although all study participants initially scored within the normal range on the MMSE, for a subset of individuals, performance declined over the five-year study period. We investigated whether baseline pTau-217, initial C3 performance, or a combination of both could predict which individuals would decline on the MMSE. Since most participants remained above the MMSE impairment threshold initially, we began predictions after three years. Demographics alone or with pTau-217 could not predict which individuals would progress to an MMSE score below 25 (Demographics: LOO AUC: 0.545 CI: 0.466 – 0. 696, pTau-217: LOO AUC: 0.631 CI: 0.490 – 0. 772). However, the model with C3 scores significantly improved predictive accuracy (Fig 3A, top, LOO AUC: 0.668 CI: 0.532 – 0. 797). Notably, the model that included both pTau-217 and C3 scores demonstrated the highest predictive value for identifying decliners (LOO AUC: 0.720 CI: 0.601 – 0. 797829).

**Figure 3:**
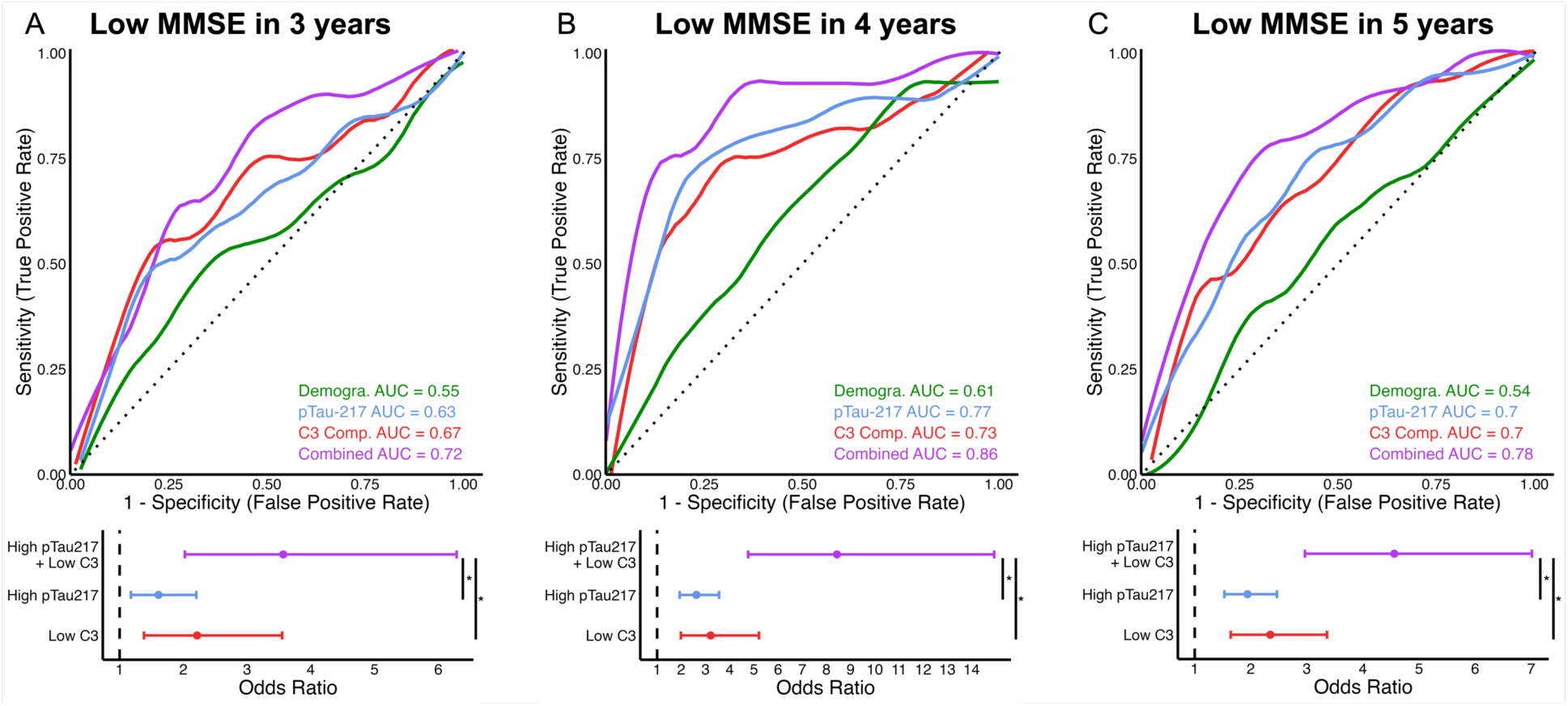
Integrating plasma pTau-217 and C3 enhances prediction of individuals who progress to clinical impairment (< 25) on the MMSE. Comparison of models using demographics alone (green), demographics plus pTau-217 (blue), C3 composite (red), or both pTau-217 and C3 (purple) for predicting low performers and future cognitive decline. (Top) ROC curves illustrating the predictive value of each model. (Bottom) Odds ratios for future impairment on the MMSE based on low memory (red), high pTau-217 (blue), or both (purple). The combined model incorporating both measures was the strongest predictor of individuals who would progress to clinical impairment on the MMSE over the next A) three, B) four, or C) five years, achieving the highest AUCs (top) and demonstrating that individuals with both high pTau-217 and low C3 performance at baseline had the highest odds ratio for progressing to an impaired MMSE (bottom).

This pattern remained consistent when predicting those who would fall below the critical MMSE threshold of 25 over four (Fig 3B, top, Demographics: LOO AUC: 0.607 CI: 0.481 – 0. 711, pTau-217: LOO AUC: 0.769, CI: 0.659 – 0.874, C3: LOO AUC: 0.733, CI: 0.619 – 0.840, Combined: LOO AUC: 0.859, CI: 0.772 – 0.928 or five years (Fig 3C, top, Demographics: LOO AUC: 0.536, CI: 0.476 – 0. 625, pTau-217: LOO AUC: 0.704, CI: 0.621 – 0.780, C3: LOO AUC: 0.698, CI: 0.620 – 0.777, Combined: LOO AUC: 0.780, CI: 0.702 – 0.851). Together, these findings further support the utility of combining pTau-217 and C3 scores to identify individuals at high risk for cognitive decline.

Next, we examined the odds of MMSE progression based on whether individuals had elevated pTau-217, low C3 scores, or both risk factors at baseline. At 144 weeks (∼2.8 years), elevated pTau-217 was associated with a 61% increased risk of progression (Fig 3A, bottom, OR = 1.610, CI = 1.177–2.204), while low memory performance was associated with a 122% increased risk (OR = 2.220, CI = 1.402–3.517). These two risk factors did not differ statistically (Z = −1.103, p = 0.270). However, individuals with both high pTau-217 and low memory had a 257% increased risk of progression (OR = 3.568, CI = 2.024–6.288), which was significantly higher than pTau-217 alone (Combined vs. pTau-217: Z = 2.241, p = 0.016) but not significantly different from low memory alone (Combined vs. Low memory: Z = 1.266, p = 0.205). This suggests that individuals with both elevated pTau-217 and subtle memory deficits are at the highest risk of progression on the MMSE.

At 192 weeks (∼4 years), we observed a similar pattern. Elevated pTau-217 was associated with a 162% increased risk of progression (Fig 3B, bottom, OR = 2.625, CI = 1.934–3.561), while low memory was associated with a 221% increased risk (OR = 3.214, CI = 1.983–5.211). Having both risk factors dramatically increased the odds of progression, with an 8.44-fold higher risk (OR = 8.436, CI = 4.764–14.938), which was significantly greater than having either high pTau-217 (Z = 3.532, p = 0.0004) or low memory alone (Z = 2.528, p = 0.011). No significant difference was found between pTau-217 and low memory alone (Z = −0.695, p = 0.487). Again, this suggests that those with subtle memory deficits along with elevated pTau-217 are particularly at risk for future cognitive decline.

Lastly, we evaluated the predictive value of pTau-217 and memory performance for MMSE decline over 240 weeks (∼4.6 years). Again, having both high pTau-217 and low memory was associated with the highest risk of progression, with a 356% increased risk (Fig 3C, bottom, OR = 4.559, CI = 2.965–7.011), significantly greater than either risk factor alone (pTau-217: OR = 1.942, CI = 1.529–2.466; low memory: OR = 2.348, CI = 1.642–3.359). Pairwise comparisons showed that the combined model was significantly more predictive than pTau-217 alone (Z = 3.399, p = 0.0007) or low memory alone (Z = 2.323, p = 0.020), with no significant difference between pTau-217 and low memory alone (Z = −0.866, p = 0.387). Together, these results further support that combining pTau-217 and memory performance improves the prediction of future decline on the MMSE.

### 3.5. Accelerated cognitive decline in individuals with both high pTau-217 and low memory performance

The findings above suggest that individuals with both elevated pTau-217 and low performance on the digital cognitive battery at baseline experienced more rapid cognitive decline. To directly investigate this, we categorized participants into four groups based on baseline pTau-217 levels and memory performance. Since, by definition, all participants had elevated pTau-217, we distinguished those with pTau-217 levels more than one standard deviation above the group mean (pTau217*) from those with pTau-217 levels at or below one standard deviation above the mean (pTau217_o_). Memory performance was classified as either normal (Mem_o_) or subtly impaired (Mem*) based on whether an individual’s memory score was more than one standard deviation below the group mean. This resulted in four groups: pTau217_o_Mem_o_ (pTau-217 within group average, no memory deficits), pTau217*Mem_o_ (pTau-217 more than one standard deviation above the group mean, no memory deficits), pTau217_o_Mem* (pTau-217 within group average, subtle memory deficits), and pTau217*Mem* (pTau-217 more than one standard deviation above the group mean, subtle memory deficits). To determine whether one of these groups declined faster, we first regressed out the effects of age, sex, and education on PACC and MMSE performance. We then examined longitudinal changes in these cognitive measures across the four groups.

We first examined PACC performance over five years across the four groups. Using linear mixed effects modeling, we found that relative to the pTau217_o_Mem_o_ group, all other groups showed significantly greater rates of decline (Fig 4A, Linear mixed-effect model: pTau217*Mem*: β = −0.00698, SE = 0.00049, t = −14.153, p < 0.001; pTau217*Mem_o_: β = - 0.00415, SE = 0.00028, t = −14.812, p < 0.001; pTau217_o_Mem*: β = −0.00251, SE = 0.00022, t = −11.594, p < 0.001). Specifically, individuals classified as pTau217*Mem* exhibited the most rapid cognitive decline compared to all other groups (vs. pTau217_o_Mem_o_: z = 6.278, p < 0.001; vs. pTau217_o_Mem*: z = 4.410, p < 0.001; vs. pTau217*Mem_o_: z = 2.486, p = 0.0129). Additionally, pTau217*Mem_o_ declined significantly faster than pTau217_o_Mem_o_ (z = 5.736, p < 0.001) and pTau217_o_Mem* (z = 2.657, p = 0.0079). Together, these findings suggest that individuals with both elevated pTau-217 and low memory performance at baseline are at the highest risk for accelerated cognitive decline, while either factor alone is associated with a moderate but significant increase in risk.

**Figure 4:**
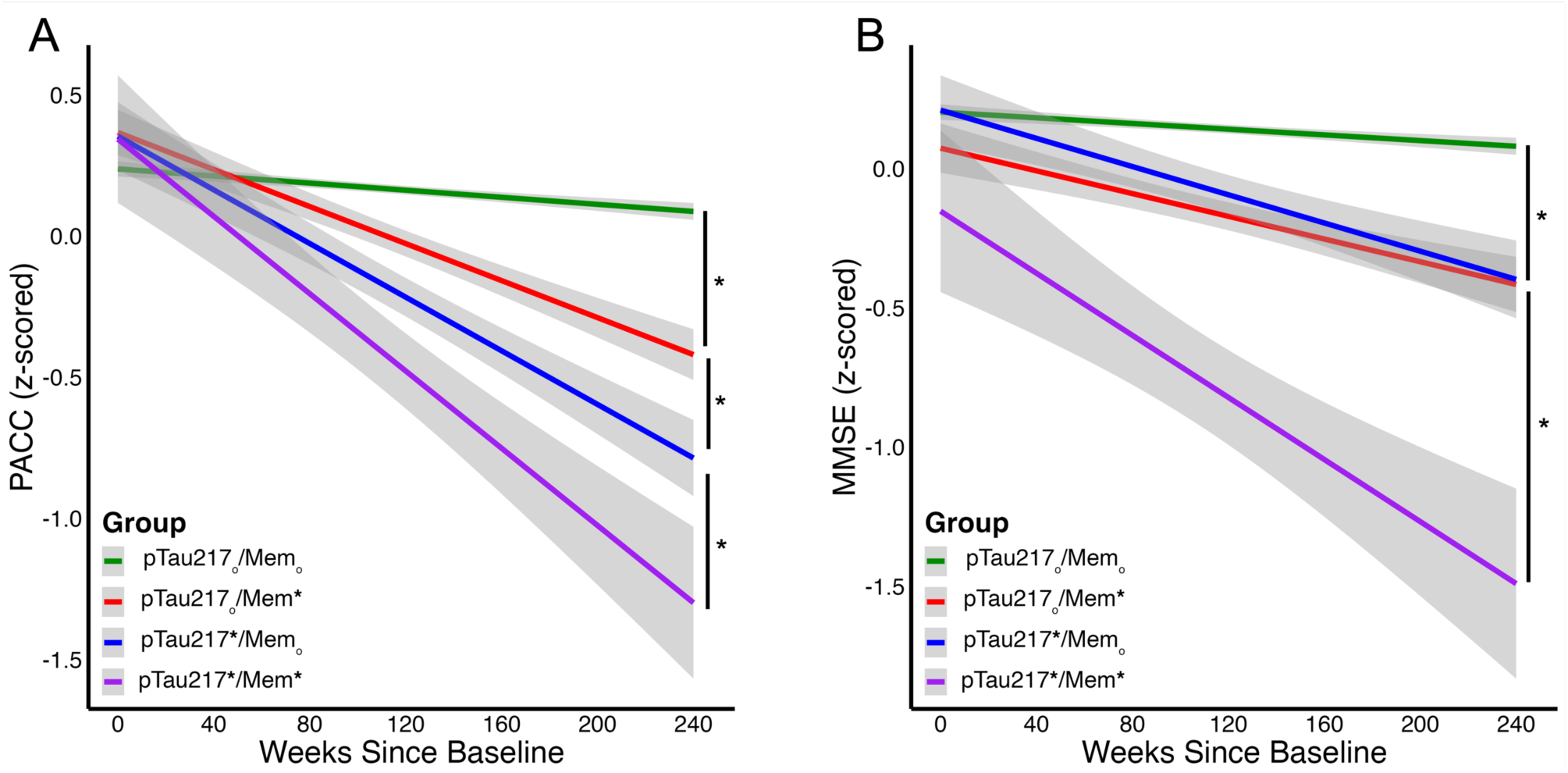
Longitudinal Trajectories Over 240 Weeks on the PACC and MMSE. (A) Individuals in the pTau217*Mem* group exhibited the most rapid decline on the PACC compared to the other three groups. pTau217*Mem_o_ showed the second fastest decline, followed by pTau217_o_Mem* which declined more rapidly than pTau217_o_Mem_o_. (B) On the MMSE, pTau217*Mem* individuals again exhibited the steepest decline. pTau217*Mem_o_ and pTau217_o_Mem* declined at similar rates, with both declining faster than pTau217_o_Mem_o_.

We next examined MMSE performance over five years across the four groups to determine whether baseline pTau-217 and memory performance predicted differential rates of cognitive decline. As with PACC, individuals in the pTau217*Mem* group exhibited the most rapid decline, followed by pTau217*Mem_o_ and pTau217_o_Mem*, while the pTau217_o_Mem_o_ group demonstrated the slowest decline. Specifically, a linear mixed-effects model revealed significant interactions between time and group, indicating distinct rates of decline among the four groups (Fig 4B, pTau217*Mem*: β = −0.00525, SE = 0.00054, t = −9.806, p < 0.001; pTau217*Mem_o_: β = −0.00201, SE = 0.00031, t = −6.527, p < 0.001; pTau217_o_Mem*: β = −0.00160, SE = 0.00024, t = −6.711, p < 0.001). Pairwise comparisons confirmed that pTau217*Mem* declined significantly faster than all other groups (vs. pTau217_o_Mem_o_: z = 8.767, p < 0.001; vs. pTau217_o_Mem*: z = 5.483, p < 0.001; vs. pTau217*Mem_o_: z = 5.911, p < 0.001). Additionally, pTau217*Mem_o_ showed a significantly steeper decline than pTau217_o_Mem_o_ (z = 3.398, p = 0.0007), while pTau217_o_Mem* also declined more rapidly than pTau217_o_Mem_o_ (z = 6.182, p < 0.001). However, no significant difference was observed between pTau21*Mem_o_ and pTau217_o_Mem* (z = −1.179, p = 0.2384), suggesting that either high pTau-217 or low memory performance alone leads to similar rates of decline, but having both greatly accelerates progression. These findings align with PACC results, further supporting that individuals with both elevated pTau-217 and poor memory performance at baseline experience the most rapid cognitive decline, whereas either factor alone confers a moderate but significant risk.

## 4. Discussion

Advances in plasma biomarkers have significantly improved our ability to readily detect AD pathology, including amyloid and tau, long before the onset of cognitive symptoms. However, the presence of these pathologies does not guarantee imminent cognitive decline; in many cases, amyloid and tau accumulate for decades before symptoms emerge, and some individuals with elevated AD biomarkers never experience measurable decline^7,32^. This uncertainty complicates decisions about when and how to intervene. To address this, we need additional low-burden, scalable tools that can refine risk stratification and provide a clearer timeline for cognitive decline. Digital cognitive assessments have emerged as a promising solution, yet their utility in complementing plasma biomarkers to identify those at highest risk remains underexplored. Here, we combined plasma pTau-217 with a composite of hippocampal memory tasks from a digital cognitive assessment in nearly 1,000 cognitively unimpaired, amyloid-positive older adults. Our findings demonstrate that integrating pTau-217 with a brief memory battery significantly enhances the ability to predict cognitive decline over the next five years. Specifically, individuals with both elevated pTau-217 and lower memory scores were at the greatest risk for future decline on both the PACC and MMSE. These results underscore the importance of incorporating both biological markers of AD pathology and hippocampal memory assessments as a more precise screening approach for identifying individuals most vulnerable to cognitive decline.

### 4.1. Integrating pTau-217 and the C3 composite for identifying individuals at risk for cognitive decline

A critical question in AD research is whether integrating plasma and cognitive biomarkers improve the prediction of future cognitive decline. Identifying individuals at high risk for decline is essential for expanding the therapeutic window for currently approved treatments and optimizing clinical trial recruitment. While prior research has established that AD biomarkers, such as amyloid, are predictive of future decline, it remains unclear whether incorporating cognitive assessments can enhance predictive sensitivity and specificity^33,34^.

Consistent with previous findings, we demonstrated that both plasma pTau-217 and memory performance were individually predictive of future decline on the PACC, with each measure contributing unique value to the models^20,21,35,36^. Further, the combined models with both measures had increased sensitivity compared to models with only one of the measures. Importantly, all analyses controlled for baseline PACC performance, ensuring that our findings were not merely driven by lower baseline cognitive scores. Our results strongly support the integration of both plasma and cognitive measures for AD detection and screening, demonstrating that a combined approach improves the prediction of cognitive decline and enhances risk stratification for clinical and therapeutic interventions.

### 4.2. Predicting decliners differs by APOE4 carrier status

Not all individuals are at equal risk for AD, with both sex and APOE4 carriership being well-established risk factors^30,37–39^. Specifically, females and APOE4 carriers face a higher likelihood of developing AD. Therefore, it is essential to validate screening tools across different populations to determine whether they perform equivalently or require adjustment based on demographic or genetic risk factors. We found that combining pTau-217 and memory performance provided comparable predictive value in males and females, both at baseline and for predicting decline over two and five years. However, while these measures were equally effective at differentiating high and low PACC performers at baseline regardless of APOE4 status, their ability to predict future decline varied. Our models performed significantly better in APOE4 noncarriers, where combining pTau-217 and memory scores achieved an AUC of nearly 0.90. In contrast, the models performed less effectively in APOE4 carriers, suggesting that APOE4 status influences the predictive utility of these measures.

APOE4 carriers are at higher risk for AD yet do not benefit as much from current treatments^40,41^. Additionally, their cognitive trajectories differ from noncarriers, with more rapid and severe hippocampal memory deficits that emerge earlier in the disease process^42–44^. In line with this, we observed reduced predictive accuracy for cognitive decline in APOE4 carriers, suggesting fundamental differences in disease pathophysiology and progression^37^. This highlights the need for further investigation into whether distinct biomarkers or cognitive measures may be required to improve risk stratification and prediction in APOE4 carriers.

### 4.3. Hippocampal memory tasks as valuable digital cognitive assessments

The use of digital cognitive tasks to identify individuals at high risk for future cognitive decline has gained increasing attention. Notably, studies have shown that remote cognitive assessments can reliably predict progression on standardized clinical measures^13,20,45^. However, not all cognitive tasks are equally effective in this regard. A consistent finding across studies is that tasks targeting hippocampal integrity are the most promising, likely due to the hippocampus being one of the earliest regions affected by AD^18,21,46^.

In this study, we incorporated three hippocampal-dependent memory tasks into our composite measure, each designed to tax pattern separation, a critical computation of the hippocampus^47–49^. The BPS-O, now widely known as the MST, was explicitly designed to assess pattern separation by requiring individuals to distinguish highly similar but not identical stimuli^25^. The OCL, adapted from the MST, requires participants to remember details about playing cards while managing competing interference from similar cards with overlapping features, thereby taxing hippocampal pattern separation mechanisms^21,50^. Lastly, the FNAME task assesses face-name associations, and given that participants must learn 12 unique pairs, successful performance likely relies on hippocampal pattern separation to differentiate and encode these associations^51–53^. These findings align with growing evidence that tasks taxing hippocampal pattern separation are early indicators of AD pathology and strong predictors of future cognitive decline^20,21,46,54^. Our results reinforce the importance of integrating hippocampal-dependent memory assessments into digital cognitive screening tools to improve early detection of at-risk individuals.

### 4.4. Clinical Utility of integrating blood biomarkers and digital cognitive testing

There is an urgent need for accessible, scalable tools to screen for AD in clinical settings, particularly in primary care, where early intervention could have the greatest impact. Blood biomarkers and digital cognitive testing are strong candidates for this purpose, as plasma collection is minimally invasive and widely available, and digital cognitive assessments can be administered remotely and autonomously at scale^12,55,56^. Our findings demonstrate the potential of these measures for identifying individuals at risk for future cognitive decline. However, critical steps remain to establish how they can be effectively implemented in real-world clinical practice.

A key challenge is that primary care providers do not have access to gold-standard neuropsychological testing, such as the PACC, and instead rely on global screening tools like the MMSE^57–59^. Here, we show that incorporating plasma pTau-217 and digital memory assessments improves the ability to predict who will decline on the MMSE over time. This suggests that a combined biomarker and cognitive screening approach could be integrated into routine clinical workflows to help identify individuals at the highest risk for cognitive decline, guiding referrals for further evaluation or early therapeutic intervention. For this approach to be clinically

actionable, future work must determine optimal cutoff points for risk classification, the most effective cognitive tasks for remote assessment, and whether additional plasma biomarkers, such as NfL or GFAP, could further strengthen predictive accuracy. Additionally, longitudinal studies are needed to assess whether this screening strategy improves patient outcomes by enabling earlier intervention and better treatment allocation.

### 4.5. Conclusion

In this study, we investigated whether integrating plasma pTau-217 with a digital cognitive assessment could improve the prediction of future cognitive decline in amyloid-positive, cognitively unimpaired older adults. Our findings demonstrate that these measures complement one another in identifying individuals at the highest risk for cognitive decline. Specifically, those with elevated pTau-217 and subtle memory deficits exhibited the most rapid cognitive decline over five years. These results highlight the potential of combining plasma biomarkers with digital cognitive assessments as low-burden, scalable tools for AD screening. We propose that this approach could be seamlessly integrated into clinical settings, enabling earlier and more precise risk stratification. By facilitating early detection, this framework has direct implications for clinical decision-making, patient monitoring, and timely therapeutic intervention, ultimately improving outcomes for those at risk for AD.

## Data Availability

All data produced in the present study are available upon reasonable request to the authors

https://www.a4studydata.org/

## Acknowledgements

The authors thank the site principal investigators of the A4 study, along with their staff, participants, and study partners. We also appreciate the Alzheimer Therapeutic Research Institute (ATRI) and the Alzheimer’s Clinical Trial Consortium for providing access to this data.

## Consent Statement

All human subjects provided informed consent.

